# IMPACT OF COVID-19 ON EDUCATION: LATEST TRENDS IN K-12 EDUCATION IN THE WORLD

**DOI:** 10.1101/2024.12.26.24319675

**Authors:** Rizwan Shoukat, Qibing Huang, Khuda Bakhsh

## Abstract

The Covid-19 has closed the whole world including the schools and colleges. This situation never thought before this in the world. Over billions of K-12 students are out of educational institutions due to this novel virus. We cannot put this risk into the future of students. Therefore, this study was conducted to explore the latest trends and practices of educational institutions which are useful during and after the Covid-19. The investigation was a mixed method (quantitative and qualitative) study which focused on questionnaires and interviews of the participants for the data gathering. A sample of 170 respondents in which 10 heads of secondary schools, 10 university professors, 50 K-12 teachers and 100 K-12 students were selected for the collection of quantitative data. However, the sample size in the collection of qualitative data was not fixed. The data collection was stopped when the emerging of new data themes stopped and the final sample of qualitative data collection was 23 participants. The triangulation technique was used to cross check the results and validation of data. The study found that the online education properly planned by the administration in the modern K-12 education in the world is the best solution during the Covid-19.

## 1. Introduction

The corona virus is the burning and most serious issue of the world. This is the only issue at the present time, which has put the whole world in trouble. The corona virus is the family of virus that may cause the range of illness from the common cold to severe disease like MERS-CoV and SARS-CoV. The Covid- 19 is the name of the disease caused by the corona virus. The Covid-19 started from the Wuhan city of China and has covered the whole world in a few months. Initially, it was novel and now it is severe acute respiratory syndrome coronavirus-2 [1]. The mortality rate as reported by [2] in February 2020 was 2.08% in China, and many confirmed cases were reported in different other cities of China including Hong Kong and Macao.

The Covid-19, whose pivot was China, spread rapidly in the other countries of the world. Within a few weeks [3], it covered 212 countries and the territories (almost all countries) of the world [4]. The major reason of the spread was travelling history of the patients outside China for the purpose of trade and business. The countries which are suffered greatly from the virus are USA, India, Brazil, Russia, UK, Turkey, Argentina, Italy, France and Iran, Pakistan is also a badly effected country from the Covid-19. Total 3,714,567 deaths were recorded on May 31, 2021 in the world.

The number of daily corona virus cases and total number of deaths on May 31, 2021 in the most affected countries of the world are reported in Table 1.

**Table 1.** Total cases and total deaths in the most affected countries.

| Sr. | Country | Total Cases | Total Deaths |
| --- | --- | --- | --- |
| 1 | India | 28173655 | 348277 |
| 2 | Brazil | 16590926 | 462966 |
| 3 | Russia | 5071917 | 121501 |
| 4 | UK | 4481973 | 127818 |
| 5 | Turkey | 5249404 | 47527 |
| 6 | Argentina | 3781784 | 78093 |
| 7 | Italy | 4217349 | 126141 |
| 8 | USA | 34039750 | 610323 |
| 9 | France | 567324 | 109533 |
| 10 | Iran | 2913136 | 80156 |

Source: Worldmeter.

### Impact of Covid-19 on Education

These are very difficult times for the parents, teachers and the students. All the stakeholders of education are worried for this pandemic. The months during which the coronavirus spread is an important period for the students, because most of the examinations are held in this period. Due to the pandemic the education system in the world has badly disturbed [5]. Cambridge Assessment International Education cancelled all the local and international examinations due to this pandemic [6]. The over billion students are out of school in the world [7]. This Covid-19 is impacting 90% of the world’ total enrollment of the students [8-9]. This massive disturbance is likely to increase the dropout rate of the students of educational institutions [10].

### Latest Trends and Practices in K-12 Education

According to Hanover research [11], the focus in the 21st century is on the use of technology in education in an innovative way. The teachers should use the modern ways of teaching using technologies. The mobile devices can be used in teaching learning process. The mobiles are very effective tools in the K-12 education because these are used by the younger children frequently. The teachers should use the appropriate technology in their classrooms to the students and for the development of lessons [12]. The technology develops more specific skills in the learners [7]. The technology gives the empowerment to the Adolescents [13]. It is a tool that can best be utilized to unlock creativity among the students [14] and it is very useful for educating the K-12 children. Students want the delivery of courses through E-learning system. The first most recently used trend among five trends which are used in the K-12 education is the technology integration into the classroom [15]. Therefore, it is an established fact that the foremost and the burning new trends of K-12 education is the incorporation of technology in the classroom.

As it is a fact that all the schools and colleges are physically closed in the world. Over billion students are out of educational institutions during Covid-19 [3]. 1.723 billion (Approx.) students have been exaggerated due to Covid-19 spread and closures of schools in the world [16]. We cannot wait more and we have to search the alternate pathways for the education [9]. The researchers in the world are struggling to rehabilitate and restore the conditions of education through the latest trends and approaches throughout the Globe. While the pandemic put all the fields in trouble, the education sector has no exception and the need being felt to provide the education through other means. The priorities of education have been changed in the corona pandemic. In the battle with Covid-19, we have to defeat the Covid-19 in the field of education. We cannot put into risk the future of our bil-lions of students in Pakistan. This emerging and burning field of research regarding coronavirus is in the infancy stage. This pandemic compelled the educators to think rationally about the online education in the world [17]. All the institutions are closed and this epidemic is so serious that none has seen this in the history of mankind. Schleicher, head of education at the organization for economic co- operation development (OECD) said that we are thinking for education solutions which have never been thought in the past [17]. This study therefore aims to explore the latest trends and practices in the K-12 education in the world and tries answering the following research questions (RQs):

RQ1: What is the present position of K-12 education in the world during Covid-19?

RQ2: To what extent is Corona-virus impacting modes of teaching and learning?

RQ3: What is the best solution for the K-12 education during Covid-19?

RQ4: How the teachers can be prepared for the online education in the world?

RQ5: What resources are needed for the K-12 online education?

RQ6: Which is the best software for online K-12 classes during Covid-19?

RQ7: How the examinations can be arranged for K-12 during Covid-19?

RQ8: What are the latest trends and practices in K-12 Education in the world?

## 2 Material and Methods

### 2-1 Research Design

The present investigation was a mixed method (quantitative and qualitative) study, focusing on the questionnaire and interviews of the stakeholders for data gathering. The triangulation technique was used to cross check the results and the data validation procedures of the study from two different data collection instruments; questionnaire and interviews. In the triangulation technique, two different data collection instruments are used [18]. The triangulation is the best way to merge the findings from two different tools to assess their validity [19-20].

### 2-2 Population and Sampling

The population of the present study was all the stakeholders of the education, including the heads of secondary schools, university professors, K-12 teachers and the K-12 students. A sample of 170 respondents in which 10 heads of secondary schools, 10 university professors, 50 K-12 teachers and 100 K-12 students were selected for the collection of quantitative data. However, the sample size in the collection of qualitative data was not fixed. The data collection was stopped when the emerging of new data themes stopped. In this way the sample size of the qualitative data collection was 23 participants.

### 2-3 Data Collection Instrument and Procedures

The interviews and the questionnaire were used as the data collection instruments. The questionnaire consisted of 15 questions which were Likert type, whereas the interview protocol comprised of 7 open ended questions to be answered by the stakeholders. The data were collected during the months of March-April, 2021. As all the respondents were at home and there was a complete lockdown in the region, therefore it was not possible to collect the data physically through visits. Therefore, the data were collected through emails and interviews. The ethics of the research were given the priority. Keeping in view the ethics, interview time was first taken from the respondents, so that it could not disturb the routine matters of the interviewee. The data collection started at the end of March 2021 and lasted till the end of April 2021. Total 23 interviews were recorded. The questions in the interview were all open ended questions. The time of the interview for the individual participant was 10 minutes to 15 minutes, with an average of a 12-minutes interview. The data collection procedure remained continue till the repetition of the qualitative data (saturation point) from the participants was reached.

### 2-4 Validity and Reliability of Data and Findings

The validity in qualitative research means the appropriateness of the research questions, data, tools, sampling, design and the methodology [21]. The validity of data and findings was checked through the focus group interviews. In this process the series of interviews was collected from the community members which were related to education, regarding the data already collected. The collected data were found appropriate by the focus groups. The validity and reliability of the coding strategy were checked by the experts in the social sciences. It was found appropriate. The reliability of the questionnaire was checked through SPSS Cronbach’s Alpha (α) method and the score of α was 0.89, which is reliable.

### 2-5 Data Analysis

The thematic analysis is a widely used method for qualitative data analysis [22]. The skills which are used in this method are the core skills of analysis [23]. The six steps suggested by Braun & Clarke [23] in the overall qualitative analysis were used in the present study. These steps are familiarizing with the data, coding, making themes, reviewing themes, naming themes and writing report. In the familiarizing phase the data were read again and again and interviews were converted into texts, in a second step the codes were made, this step is the part of analysis of data which converts the collected data into meaningful groups [24]. There are different methods of coding and in the present investigation inductive coding technique was used. In the third step the themes were framed from the codes, which were reviewed in the fourth step and all were found appropriate. The fifth step was themes naming, these names were succinct, punchy and easy and finally the report regarding the qualitative analysis was prepared. However, the analysis of quantitative data was completed through the SPSS software. The t-test, percentage and means were used for the data analysis.

## 3 Results

### 3-1 The Quantitative Data Analysis

The Panel A of Table 2 showed the frequencies and percentages indicating that 86.4% participants agreed that the online learning is the best solution during Covid-19 for K-12 education, the majority (71.7%) were of the view that online education is approachable for developing countries. 57.6% agreed that the teachers are well trained in online learning. 97.4% participants viewed the online education as time saving.

**Table 2.**
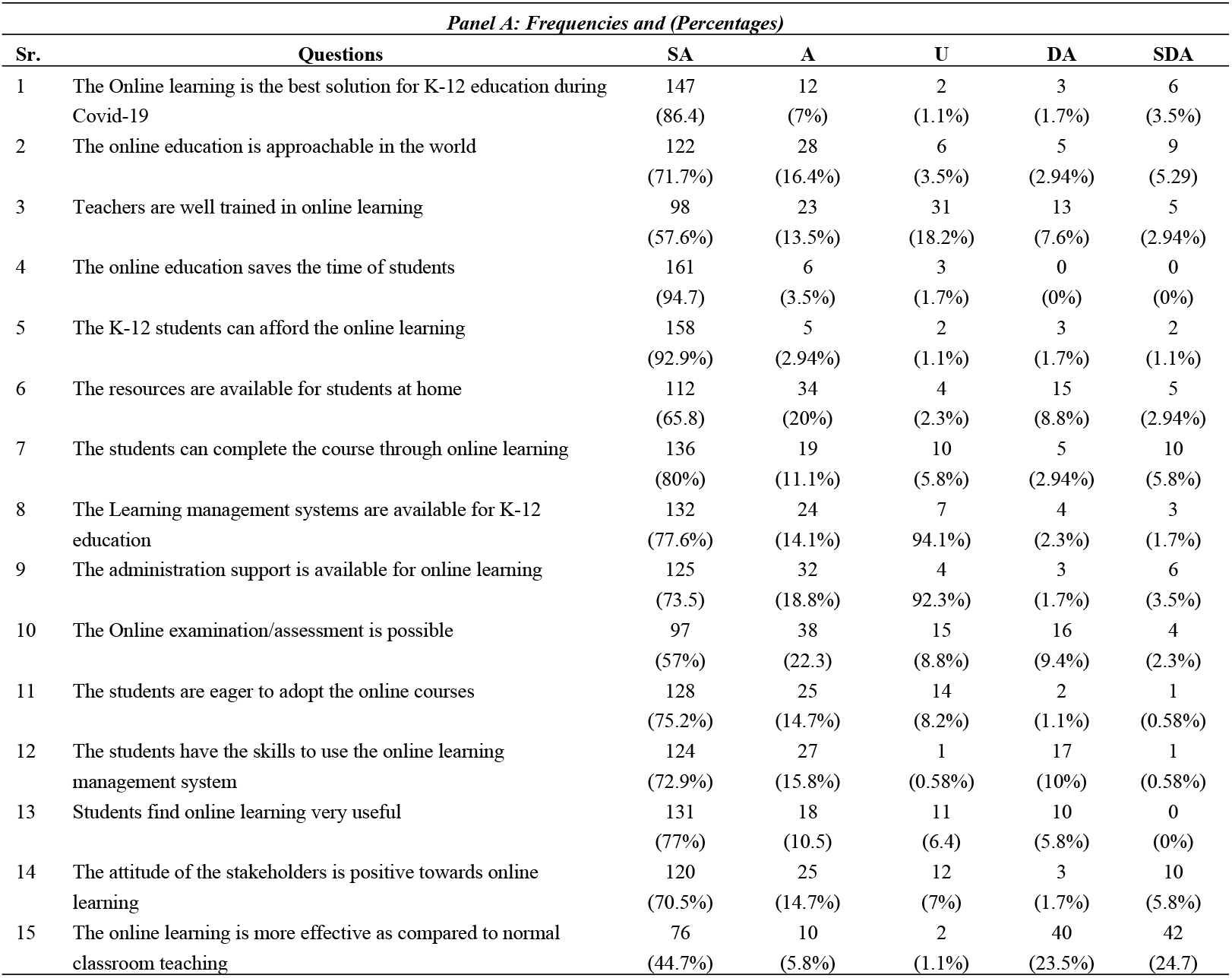
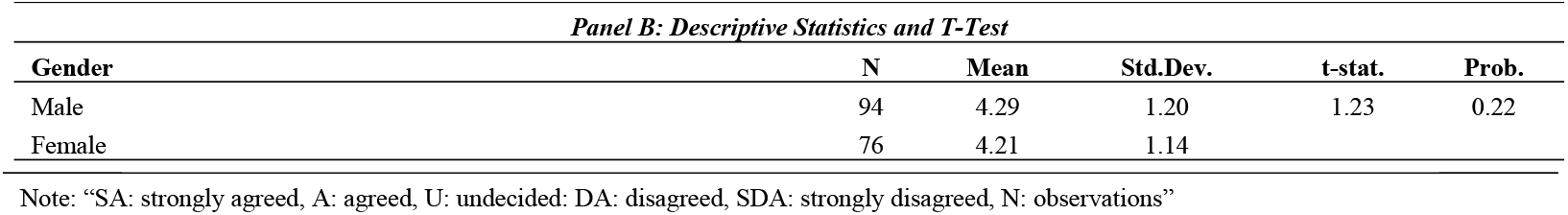
Frequencies, percentages, descriptive measures and t-test.

| *Panel A: Frequencies and (Percentages)* |  |  |  |  |  |  |
| --- | --- | --- | --- | --- | --- | --- |
| Sr. | Questions | SA | A | U | DA | SDA |
| 1 | The Online learning is the best solution for K-12 education during Covid-19 | 147<br>(86.4) | 12<br>(7%) | 2<br>(1.1%) | 3<br>(1.7%) | 6<br>(3.5%) |
| 2 | The online education is approachable in the world | 122<br>(71.7%) | 28<br>(16.4%) | 6<br>(3.5%) | 5<br>(2.94%) | 9<br>(5.29) |
| 3 | Teachers are well trained in online learning | 98<br>(57.6%) | 23<br>(13.5%) | 31<br>(18.2%) | 13<br>(7.6%) | 5<br>(2.94%) |
| 4 | The online education saves the time of students | 161<br>(94.7) | 6<br>(3.5%) | 3<br>(1.7%) | 0<br>(0%) | 0<br>(0%) |
| 5 | The K-12 students can afford the online learning | 158<br>(92.9%) | 5<br>(2.94%) | 2<br>(1.1%) | 3<br>(1.7%) | 2<br>(1.1%) |
| 6 | The resources are available for students at home | 112<br>(65.8) | 34<br>(20%) | 4<br>(2.3%) | 15<br>(8.8%) | 5<br>(2.94%) |
| 7 | The students can complete the course through online learning | 136<br>(80%) | 19<br>(11.1%) | 10<br>(5.8%) | 5<br>(2.94%) | 10<br>(5.8%) |
| 8 | The Learning management systems are available for K-12 education | 132<br>(77.6%) | 24<br>(14.1%) | 7<br>(4.1%) | 4<br>(2.3%) | 3<br>(1.7%) |
| 9 | The administration support is available for online learning | 125<br>(73.5) | 32<br>(18.8%) | 4<br>(2.3%) | 3<br>(1.7%) | 6<br>(3.5%) |
| 10 | The Online examination/assessment is possible | 97<br>(57%) | 38<br>(22.3) | 15<br>(8.8%) | 16<br>(9.4%) | 4<br>(2.3%) |
| 11 | The students are eager to adopt the online courses | 128<br>(75.2%) | 25<br>(14.7%) | 14<br>(8.2%) | 2<br>(1.1%) | 1<br>(0.58%) |
| 12 | The students have the skills to use the online learning management system | 124<br>(72.9%) | 27<br>(15.8%) | 1<br>(0.58%) | 17<br>(10%) | 1<br>(0.58%) |
| 13 | Students find online learning very useful | 131<br>(77%) | 18<br>(10.5) | 11<br>(6.4) | 10<br>(5.8%) | 0<br>(0%) |
| 14 | The attitude of the stakeholders is positive towards online learning | 120<br>(70.5%) | 25<br>(14.7%) | 12<br>(7%) | 3<br>(1.7%) | 10<br>(5.8%) |
| 15 | The online learning is more effective as compared to normal classroom teaching | 76<br>(44.7%) | 10<br>(5.8%) | 2<br>(1.1%) | 40<br>(23.5%) | 42<br>(24.7) |
| *Panel B: Descriptive Statistics and T-Test* |  |  |  |  |  |  |
| Gender | N | Mean | Std.Dev. | t-stat. | Prob. |  |
| Male | 94 | 4.29 | 1.20 | 1.23 | 0.22 |  |
| Female | 76 | 4.21 | 1.14 |  |  |  |

Note: “SA: strongly agreed, A: agreed, U: undecided: DA: disagreed, SDA: strongly disagreed, N: observations”

The K-12 students in the world can afford online learning was favored by 92.9%. 65.8% said that resources like computer, internet and webcams are available for online teaching. 80% said that K-12 students can complete the courses online. 77.6% participants were of the view that learning management systems are available. 73.5% said that administration support is available for online learning. 57% participants said that the online examinations can be done. 75.2% participants are of the view that students are willing to adopt the online classes, and this online learning is very useful and effective for them (77% strongly agreed). 70.5% participants were of the view that the stakeholder attitude is positive towards the online education and 44.7% viewed it as more effective than the real classroom teaching while 72.9% respondents were of the viewed that the students have the skills to use the online learning management system. The analysis in Panel B of Table 2 portrayed that the mean of the male and female was 4.29 and 4.21 respectively. The overall analysis depicted no difference (see Figure 1) in the views of male and female participants (t=1.23, p=0.22 > 0.05).

**Figure 1.**
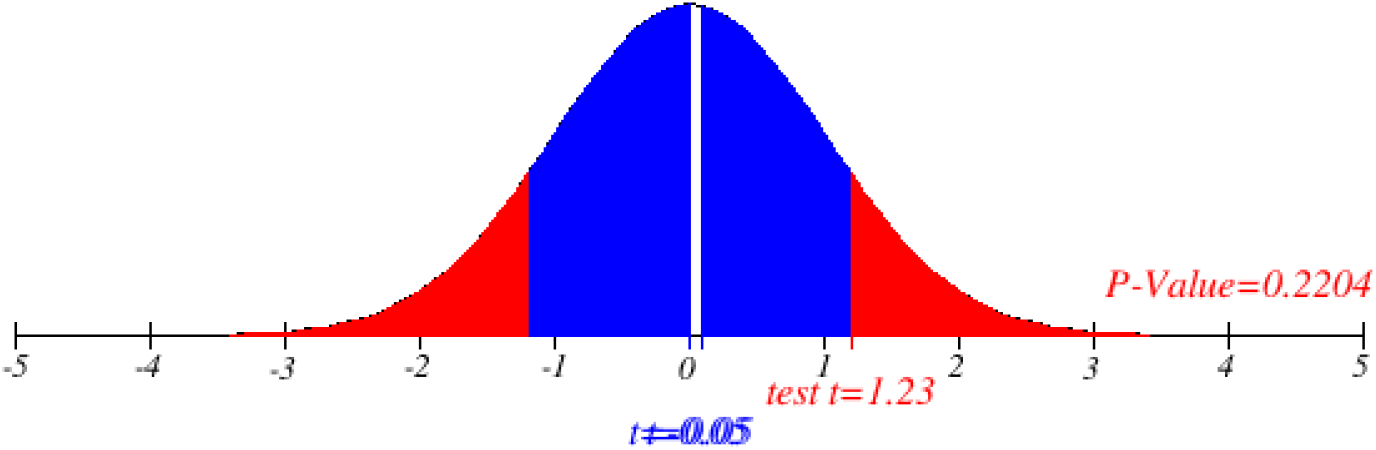
T-Test: Difference between the views of males and females.

### 3-2 The Qualitative Data Analysis

The content analysis was done in the examination of qualitative data. The Table 3 show the detailed qualitative data analysis. The interviews were transcribed into text, the transcripts were converted into codes to organize the data and to pick the themes. The analysis of the data depicted that there was completely lockdown and all the K-12 educational institutions were closed. The coronavirus had greatly impacted the modes of teaching and learning at K-12 level, as the physical classrooms were not possible, therefore the online teaching was the best solution at the K-12 level during Covid-19. The software which could best be utilized in the world was Zoom, Moodle and Google classrooms that are freely available online. For the teachers and students to run effectively the K-12 online classes there is need of some infrastructure which may include the computer with a webcam and the internet facility. The teachers need to be trained in taking the K-12 online classes. During the lockdown the training of teachers is not possible through physical appearance in the sessions, therefore online tutorials and online training managed by authorities is the best solution. The final step is the K-12 examination, that needs to be con-ducted through viva, because in this way the chances of cheating can be minimized.

**Table 3.** Selected interview responses & codes and codes & emerging themes.

| *Panel A: Selected interview responses and codes* |  |  |
| --- | --- | --- |
| Topic | Interview Response | Codes |
| Present position of K-12 education in the world | A-The Covid-19 has disturbed education system as well as examinations.<br>B- Disturbed the education system and millions of students are out of schools due to Lock down | Disturbance, Examination, Millions of Students out of school and Lockdown |
| Corona-virus impacts on modes of teaching and learning in K-12 Education | A- Due to this pandemic the students are out of schools, the need is to shift the classes on online system<br>B- Due to this pandemic the institutions are thinking for the online distance learning | Shifting of Classes to online<br>Online distance learning |
| Best solution for the education of K-12 students during Covid-19 | A- Online classes are the best, but the administrators must keep in mind the internet facility<br>B- The online learning through LMS and Virtual teaching platform is the best solution of education | Online classes,<br>Internet facility,<br>Virtual teaching platform, and Learning management system |
| Best software for K-12 online classes during Covid-19 | A- Moodle and the recorded lectures are the best solution<br>B- Zoom software and the google classroom can be used for the online classes during Covid-19 | Recorded lectures, Zoom software, Google classroom, Moodle software |
| Resources for K-12 online education | A- Computer and internet facility<br>B- The internet, webcam, and the computer | Computer, Internet facility, Webcam |
| Teachers preparation for the K-12 online education | A- Tutorials are available for learning<br>B- The teachers can be trained by management through online training | Tutorials,<br>Online Training |
| Examinations arrangement for K-12 online courses | A- Exams are possible through Viva-voce<br>B- The Proctor Track services can be taken, also the viva examinations can | Viva-voce examination<br>Proctor Track services |
|  | be conducted to avoid cheating | Avoid cheating |
|  | **Panel B: Codes and emerging themes** |  |
| **Topics** | **Codes** | **Themes** |
| Present position of K-12 education in the world | Disturbance, Examination, Millions of Students out of school and Lockdown | Lockdown, Institutions closed |
| Corona-virus impacts on modes of teaching and learning in K-12 Education | Shifting of classes to online and Online distance learning | Online teaching |
| Best solution for the education of K-12 students during Covid-19 | Online classes, Internet facility, Virtual teaching platform, Learning management system | Online education |
| Best software for K-12 online classes during Covid-19 | Recorded lectures, Zoom software, Google classroom, Moodle software | Zoom, Moodle, Google Classroom |
| Resources for K-12 online education | Computer, Internet facility, Webcam | Computer, Internet |
| Preparation for K-12 online education | Tutorials, Online Training | Training, Tutorials |
| Examinations arrangement for K-12 online courses | Viva-voce examination, Proctor Track services, Avoid cheating | Viva examination |

### 3-3 Triangulation

The multiple sources for collecting data to ensure the validity of results is triangulation [25]. In the present study the questionnaire and the interview methods were used for the triangulation. Triangulation is used in the mixed method research where the results from different sources are combined to each other [26]. Therefore, the present study’s tri-angulation indicates that there is complete lockdown and the K-12 education has been halted. The only solution is the online learning.

## 4 Discussion: Online Learning as the New Trend in K-12 Education During Covid-19

The online learning is actually based on the constructivist ideology [27-28]. In the online learning, both teachers and the students have to pay extra energy and efforts, but they give the best results [27]. This is a self-regulatory teaching in which goals are specified as the goal specification is the basic requirement [29]. The programs are developed to achieve those goals and then revisions/modifications are made keeping in view the final evaluation [30]. This trend and strategy of education is not only limited to Covid-19, rather it can be used to supplement the classroom learning in a normal classroom. This is an interactive approach/model. According to Fasso [31], the interactive approach is more effective in online teaching. Before this model the researchers failed to present the complete online learning model which could fulfil the requirements of the users, however blended learning model was floated by the researchers before this model which is the blend of classroom and e-learning [32].

The online learning model shown in Figure 2 demonstrated the program of K-12 education during the Covid-19 in the world. The first and the foremost necessity is the sup-port of administration and management. The management may then decide the training of teachers and the staff for the online delivery of education at a K-12 level. The development of skills of the teachers is compulsory for online teaching [33]. The model depicted that apart from the training of teachers-students’ awareness of computer is also necessary. The students like the delivery of courses through internet [34]. The most important point to consider here is the resources/infrastructure, including computers, net facilities and all other requirements that can be made available at the grass root level for proper functioning of K- 12 online classes. The internet is an important resource in online learning [28]. This model is antenatal with the development of the K-12 online courses because face-to-face teaching within the physical classroom is somewhat different from the online teaching, therefore, the courses must be revised which could fulfill the needs and demands of the K-12 online teaching. The online course length is sufficient in the range of 16-18 weeks [29]. Keeping in view the course development, the delivery of these courses could be made possible, which could be done through asynchronous and synchronous modes of delivery. The blend of both (synchronous and asynchronous) modes is best suitable for students during the Covid-19. In the synchronous mode, Zoom software is best which has the facility of online lectures, in which the students and teachers can see each other. This is like the Skype video call or the video conferencing. The Zoom and Moodle software have the capacity to interlink the dozens of students and teacher within an interactive online teaching. Moodle has the capacity of face-to-face learning [30-31]. The second phase of the course delivery is through google classroom. This is the asynchronous portion of online lectures which has multiple facilities like the delivery of the assignments to the students. The attachments of the course materials through e-mail, asking questions, etc. The final step of this 6-factor model is the online evaluation which is really a difficult task, because in the online teaching, papers cannot be distributed online, as the teacher does not know who one is answering at the students’ end. Therefore, the interviews and the face-to-face online question answers are suggested for the K-12 online evaluation of students. After the evaluation, if the teacher feels that there are some drawbacks or the results are not more satisfactory then the facility of review is available in which the teacher or administration may review the steps to see that where is the weakness and the same may be removed to make the model 100% effective for online teaching and evaluation.

**Figure 2.**
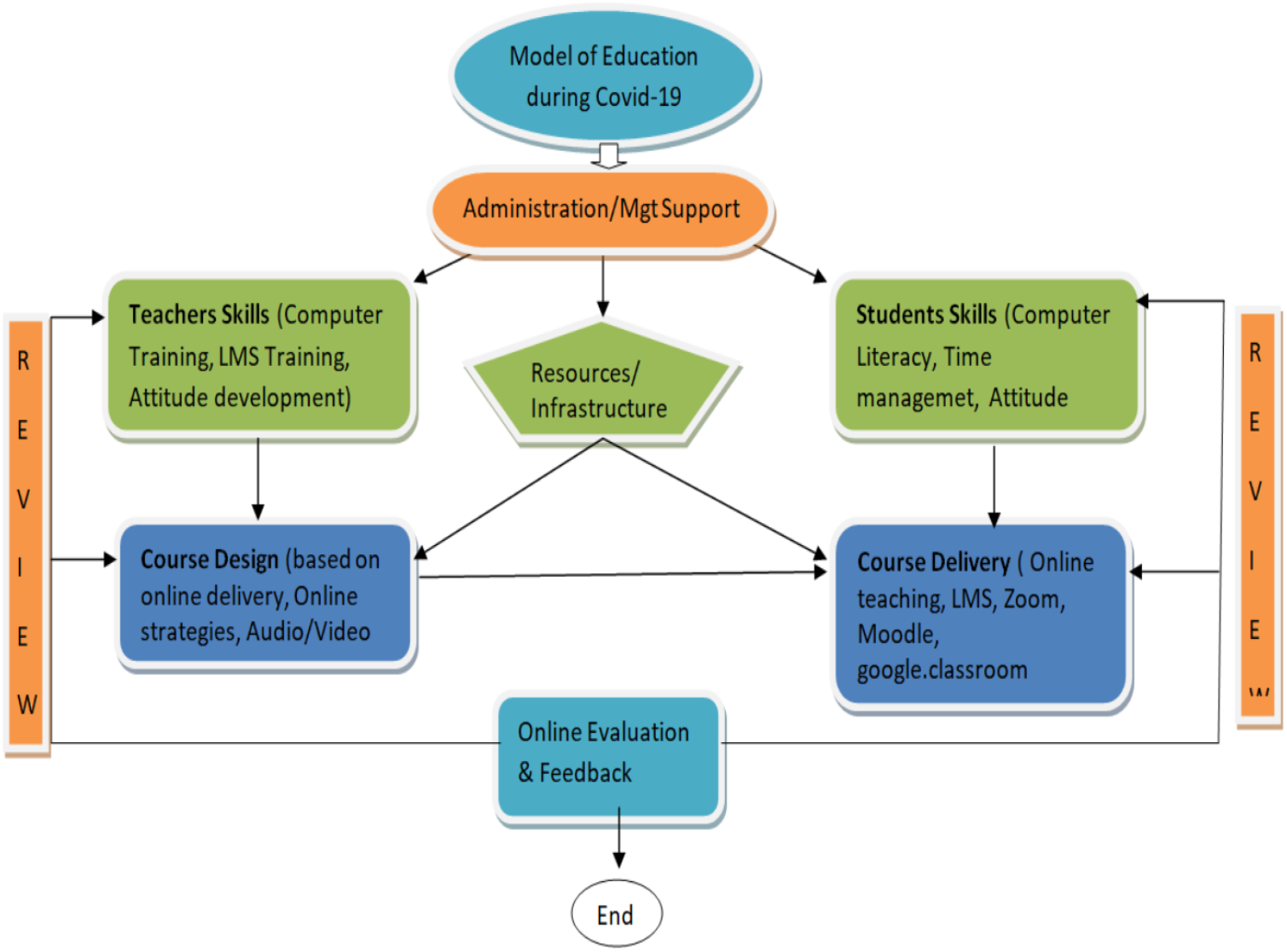
Latest trends and practices in K-12 education in the world.

## 5 Conclusion

Due to the pandemic the education system in the world has badly disturbed as the coronavirus had greatly impacted the modes of teaching and learning at K-12 level as the physical classrooms were not possible, therefore, the online teaching was the best solution at the K-12 level during Covid-19. The software which could best be utilized in the world was Zoom, Moodle and Google classrooms that are freely available online. The delivery of courses could be made possible through asynchronous and synchronous modes. The blend of both modes is best suitable for students during Covid-19. In the synchronous mode the Zoom software is best which has the facility of online lectures, this is just like the video conferencing. The Zoom and Moodle software have the capacity to interlink the dozens of students and teacher within an interactive online teaching. Moodle has the capacity of face-to-face learning. The second method of the course delivery is Google class-room which is the asynchronous portion of online lectures and has the multiple facilities like delivery of assignments to the students. Hence, it is concluded that the online education properly planned by the administration in the modern K-12 education in the world is the best solution during the Covid-19.

## 6 Declarations

### 6-1 Author Contributions

Conceptualization, MKB and SHT; methodology, MKB; software, MRU; validation, LH, NS and MRU; formal analysis, MKB; investigation, SHT; resources, MKB.; data curation, MRU; writing—original draft preparation, SHT; writing—review and editing, LH; visualization, SHT; supervision, NS; project administration, MRU. All authors have read and agreed to the published version of the manuscript.

### 6-2 Data Availability Statement

The data presented in this study are available on request from the corresponding author.

## 6-4 Acknowledgements

We thank Government College University Faisalabad, Pakistan, to use the library and internet services to complete the research article.

